# Multisite evaluation and validation of Optical Genome Mapping for prenatal genetic testing

**DOI:** 10.1101/2022.12.19.22283552

**Authors:** R.E. Stevenson, J. Liu, A. Iqbal, B. DuPont, N. Sahajpal, M. Ho, J.W. Yu, S.J. Brody, M. Ganapathi, A. Rajkovic, T. Smolarek, F. Boyar, P. Bui, A.M. Dubuc, R. Kolhe, B. Levy

## Abstract

Cytogenetic studies represent a critical component of prenatal genetic testing. Prenatal diagnostic testing of amniotic fluid, chorionic villus sampling, or more rarely, fetal cord blood, is recommended following a positive or unreportable NIPT, maternal serum screen, abnormal ultrasound or increased genetic risk based on family history. While chromosomal microarray is the recommended first-tier prenatal diagnostic test for the detection of sub-microscopic copy number variants, in practice, multiple assays are often assessed, in concert, to achieve a final diagnostic result. The use of multiple methodologies is costly, time consuming, and labor intensive.

Optical genome mapping is an emerging technique with application for prenatal diagnosis because of its ability to detect and resolve, in a single assay, all classes of pathogenic cytogenetic aberrations detectable by karyotyping, FISH, and microarray. In an effort to characterize the potential of optical genome mapping as a novel alternative to conventional testing, a multi-site, multi-operator, multi-instrument clinical research study was conducted to demonstrate its analytic validity and clinical utility. In the first phase a total of 200 specimens representing 123 unique cases demonstrated 100% concordance with standard of care methods and 100% reproducibility between sites, operators, and instruments. Analysis and interpretation of cases with incidental findings of potential clinical significance also were performed.

## Introduction

Technological innovation and subsequent adoption of non-invasive prenatal testing (NIPT) has dramatically altered the landscape of prenatal genetic screening. Today, the American College of Obstetrics and Gynecology and the American College of Medical Genetics and Genomics recommend the option of NIPT for all women, irrespective of age or risk factors (1,2). This sequence-based technology has facilitated a greater sensitivity for the detection of aneusomy of chromosomes 13, 18, and 21 and sex chromosomes compared to traditional maternal biochemical serum screening for aneuploidy (3–5). Nonetheless, diagnostic testing via chorionic villus sampling (CVS) or amniocentesis is recommended for confirmation of a positive or uninformative NIPT result. As such, NIPT is considered to be a screening test. NIPT also is utilized to screen for select microdeletions/microduplications and other rare autosomal trisomies but shows relatively poor positive predictive value (PPV) for these applications (6,7).

Diagnostic testing via chorionic villus sampling (CVS) or amniocentesis remains the recommended course of action when a definitive diagnosis is sought, when fetal anomalies are observed by ultrasound and when there is a positive family history for adverse perinatal outcomes. Chromosome G-banding analysis, commonly referred to as karyotyping (KT), has been the gold standard prenatal diagnostic test for over 50 years (8). KT offers genome-wide detection of large visible copy number variations (CNVs) as well as both balanced and unbalanced structural variants (SVs). The resolution of KT is typically considered to be ∼ ≥5-10 Mb, and SVs and CNVs below this size cutoff will be missed. These sub-microscopic imbalances are referred to as microdeletions and microduplications. Chromosomal microarray analysis (CMA) provides improved resolution for the detection of CNVs ranging from ∼50 kbp and up, depending on the probe size and placement of the array platform used (9,10). While CMA increases diagnostic yield in prenatal diagnosis (11), it should be noted that KT is still required to detect (balanced) structural rearrangements, which represent a well-described diagnostic ‘blind spot’ of CMA. CMA has become a first-tier diagnostic test at greater frequency across the globe, particularly when fetal anomalies are encountered. However, KT, FISH, MLPA and qPCR remain the tests of choice in many countries, primarily owing to financial and resource constraints (6,7,9,12).

Since numerous prenatal diagnostic options are available, consideration is given to the inherent strengths and limitations of each technology when deciding which to use. In practice, a combined approach is often employed with multiple tests ordered, either concurrently or consecutively. For example, FISH for the common aneusomies may be utilized to provide a rapid result following an abnormal screening test. A negative FISH result would prompt further testing by KT or CMA. In many cases, CMA is performed concurrently with KT and in some cases, CMA is only performed if KT is normal. A positive FISH or CMA result may still trigger KT to identify the nature of the anomaly to define potential future reproductive risks (e.g. suspected translocations in which a parent may be a carrier).

Optical genome mapping (OGM) is an emerging cytogenomic technology that offers a cost effective, genome-wide assessment of all forms of variants (SVs and CNVs) at a resolution which exceeds that of CMA (13–15). Additionally, this assay has the potential to detect certain repeat expansions, facilitating detection of additional genomic alterations which may contribute to clinical management. For example, OGM allows for the screening of the Fragile X syndrome (FXS) expanded allele (full mutations) by running a different analysis pipeline algorithm (EnFocus Fragile X) on the same sample in addition to the routine OGM whole genome analysis. Carrier testing for FXS is recommended by ACOG, ACMG and other professional societies as FXS is the most common form of inherited intellectual disability and autism (incidence of ∼ in 4,000 males and 1 in 5,000 females) (16). OGM works by fluorescent labeling of ultra-high molecular weight DNA (UHMW DNA) at specific 6-bp motif sites (CTTAAG) occurring approximately every 5 kbp throughout the genome. The labeled molecules are aligned to a reference genome to detect genomic aberrations as small as 500 bp and as large as whole chromosomal aneusomies or triploidy. Recent studies have shown that OGM has high (∼100%) concordance with traditional cytogenetic techniques in constitutional settings including prenatal and postnatal applications (13,17–19).

## Methods

The primary aim of this study was to perform a muti-site evaluation of the performance, robustness, and reproducibility of OGM compared to Standard of Care (SOC) technologies. To facilitate this goal, a standardized analysis and interpretation workflow was developed for the classification of SVs detected by OGM across multiple sites, thereby reducing the variability in SV reporting (a common challenge observed amongst the participating sites while reporting CNVs from CMA).

### Cohort design

This double-blinded observational study was approved through multiple IRBs, and the first phase included 200 de-identified independent samples (including replicates) representing 123 unique individuals (Figure 1). Remnant clinical samples were obtained from individuals who underwent invasive prenatal testing, either by CVS or amniocentesis. Samples were given anonymous aliases used in this study (i.e.: BNGOSS-xxxx). The study was specifically designed to include a variety of clinical cases with known chromosomal aberrations (e.g.: deletion, duplication, insertion, aneuploidy, inversion, translocation, and other complex variants) as well as cases reported with large regions of homozygosity (Figure 2). Cases with pathogenic/likely pathogenic sequence variants (e.g. single nucleotide variants and indels), Robertsonian translocations, balanced centromeric translocations, and mosaicism (< 20% cellular fraction) were excluded from this study.

**Figure 1.**
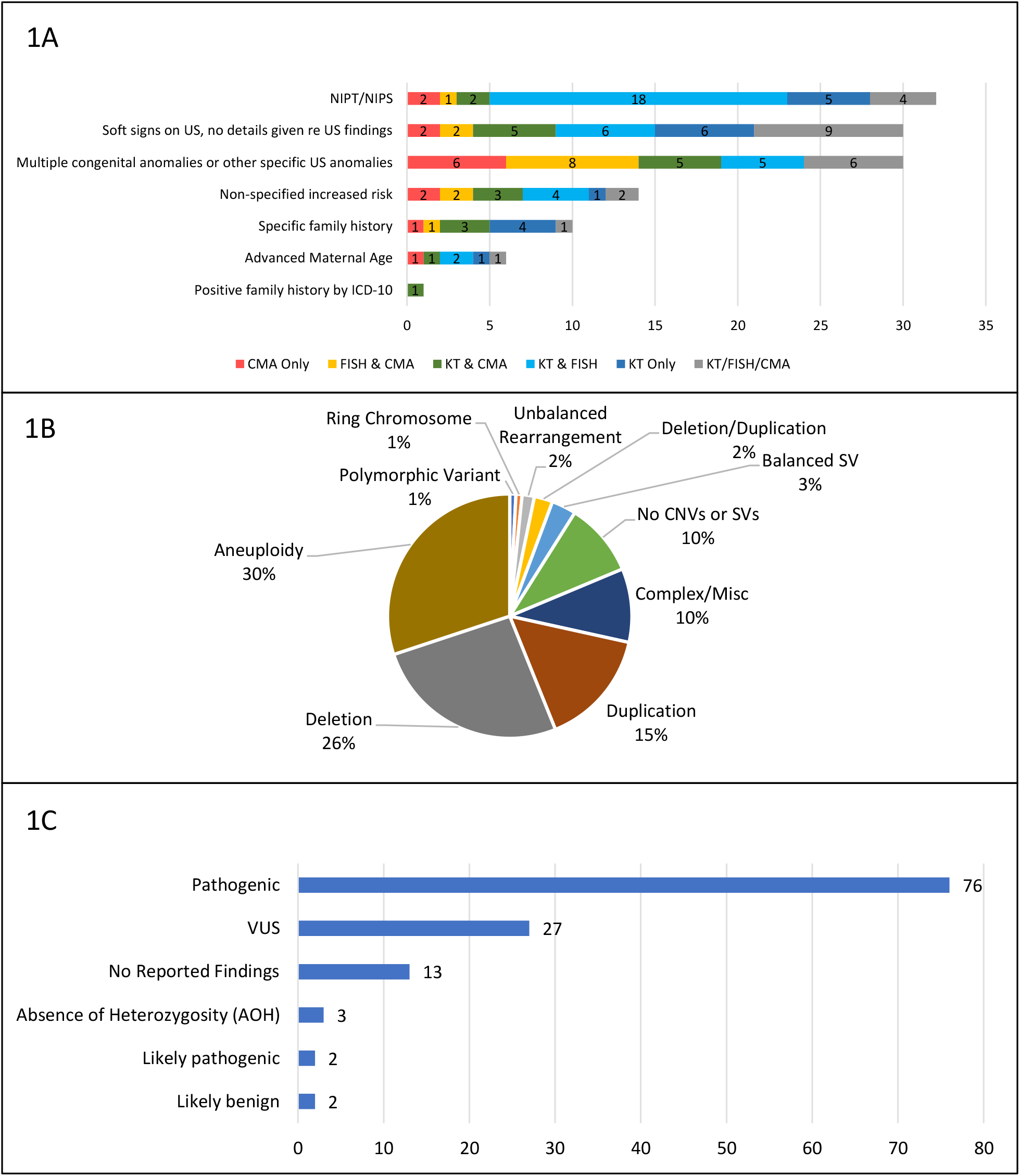
Descriptions of the 123 unique prenatal cases in this cohort. 1A) Breakdown of cases by clinical indication category and what cytogenetic SOC tests were performed. 1B) Cases by reported SOC structural variation class. 1C) Cases by ACMG guidelines classification of structural variations.

**Figure 2.**
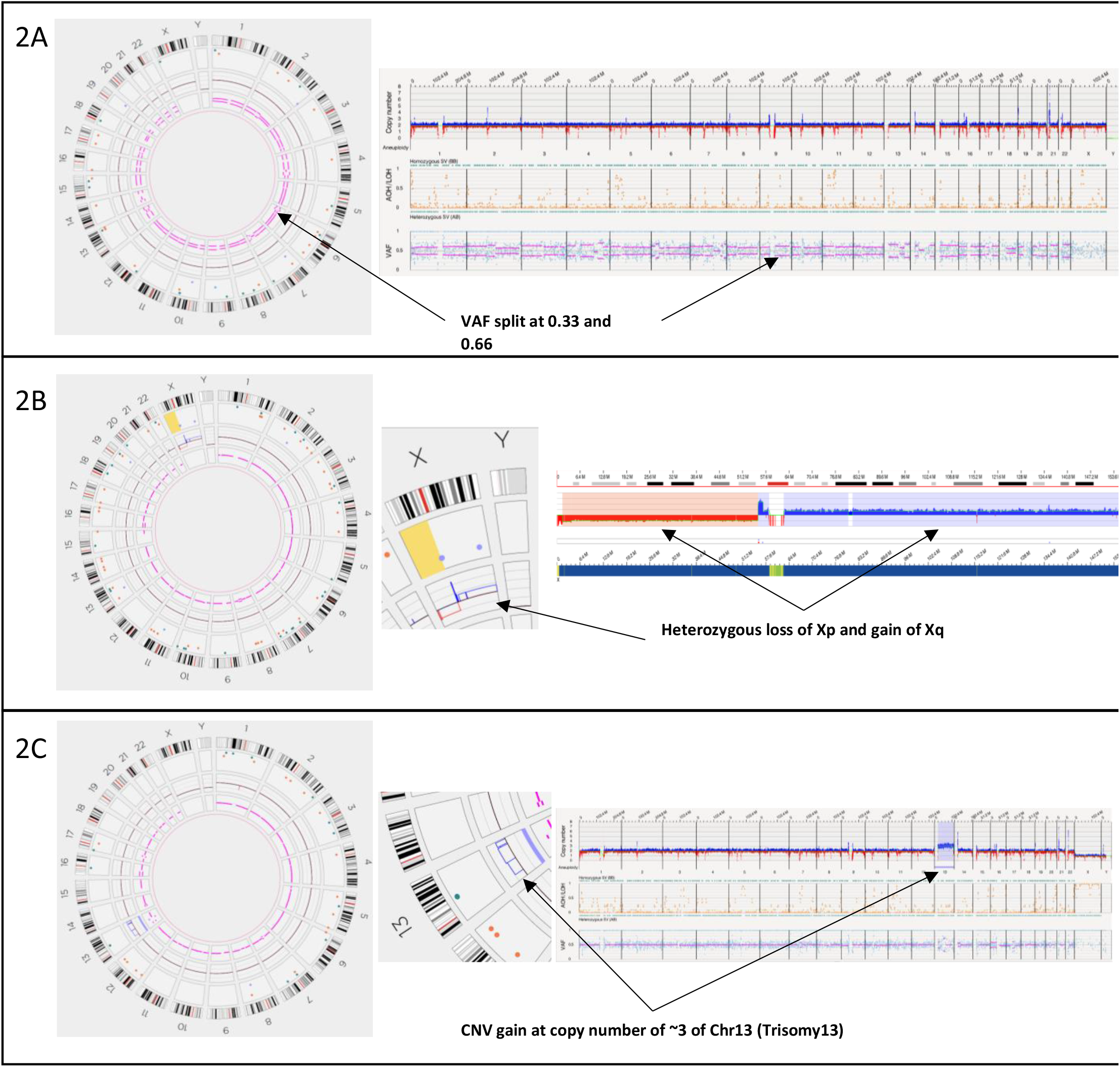
Representative examples of structural variant classes detected by OGM. 2A) Triploidy is shown in the Circos plot and whole genome view with stable genome-wide copy number but split VAF tracks at 0.33 and 0.66. 2B) Dicentric X chromosome with a copy number loss of Xp and a gain of Xq. 2C) Whole genome view showing a gain of chromosome 13 (trisomy 13).

### Sample preparation and processing

Aliquots of cryopreserved amniocytes or CVS samples were prepared at sample-contributing sites. Briefly, cells cultured in flasks were detached with trypsin-EDTA, rinsed into conical tubes and pelleted, counted, and cryopreserved in freezing medium with 5% DMSO targeting 1 – 1.5 million viable cells per sample. Samples were shipped after deidentification to a central storage location and each sample was assigned a new study identification number. The cryopreserved aliquots remained frozen until DNA isolation at a sample processing site.

UHMW DNA was isolated and labeled using Bionano Prep® SP v2 and Direct Label and Stain (DLS) kits (Bionano Genomics, San Diego, CA, USA). Cryopreserved samples were thawed and recounted, washed, pelleted from supernatant, resuspended, then digested with Proteinase K, RNase A, and lysis buffer. DNA was precipitated with isopropanol and bound to a nanobind magnetic disk, washed successively, then eluted into the elution buffer overnight. Solubilized DNA was quantitated fluorometrically, and then 750 ng was labeled at a 6-bp motif (‘CTTAAG’) using a DLS assay. Labeled DNA solution was quantitated for quality control, then loaded onto the Saphyr® chips for imaging. The fluorescently labeled DNA molecules were imaged sequentially across nanochannel arrays on the Saphyr® instrument. An 800 Gbp (effective coverage of ≥160×) throughput target was set for data collection for each sample. Three main analytical QC targets were set for passing criteria: molecule N50 (≥150 kbp) >230 kbp, map rate of molecules to the reference genome ≥70%, and effective coverage of the genome ≥160×.

***Data analysis and SV interpretation***

Genome analysis was performed using Bionano Solve™ (v3.7) software and OGM specific pipelines managed via Bionano Access™ (v1.7) software. EnFocus™Fragile X pipeline was run on all samples internally to measure the length of the genomic region corresponding to the 5’ UTR of the *FMR1* gene and that measurement was used to infer and calculate the CGG repeat for each allele found. In some cases, repeat lengths are inferred to be <1 repeat; these are considered not expanded. Additional statistics were calculated by the EnFocus™ Fragile X pipeline, such as the inferred sex of the case based on presence or absence of Y-chromosome material, an assessment of DNA molecule quality and integrity, uniformity of molecule mapping (used for CNV calling by read depth), and an assessment of stable regions in the genome (20). The stable regions are established by the manufacturer for 22 regions across the genome, one for each autosome, which do not contain any SVs in a control database (20).

Next, a genome wide analysis was performed using the Bionano Solve *de novo* assembly pipeline (21). The *de novo* assembly pipeline outputs four additional quality assessments to measure the amount of noise in each sample’s coverage profile. The statistics measured whether in the sample there was a notable elevation in 1) the correlation of variation of coverage against a theoretical baseline (using window size of 2 Mbp and 6 Mbp), 2) the correlation with label density, and 3) the correlation with known wavey coverage profiles. All samples that failed these metrics were noted.

In the *de novo* assembly pipeline, samples were compared to the human reference genome (GRCh38) to identify structural variants (SV), copy number variants (CNV), and absence of heterozygosity/regions of homozygosity (AOH/ROH). AOH/ROH segments were detected with an expected resolution of ≥25 Mbp. SVs and CNVs were first passed through a baseline set of software default filters, to remove low confidence calls and possible technical artifacts. Analysts trained at analyzing OGM data then applied a set of additional filters approved by the board-certified laboratory directors and principal investigators of this study, viz. variants ≥1.5 kbp in size, present in ≤1% controls of an OGM-specific internal SV control database (22) and overlapping or within 3 kbp of a gene (this criterion was not applied to translocation and large fusion breakpoints). Variants meeting these criteria were then curated for review by the directors and classified based on ACMG guidelines (23).

Detailed information regarding cohort design, sample processing, assay quality control, data analysis, variant interpretation, and clinical concordance/classification is available in the supplemental information (Table S1).

## Results

### Cohort description and SOC testing

Of the 200 samples (datapoints) in this cohort that underwent variant analysis, 123 were unique prenatal cases (n=114 amniotic fluid and n=9 CVS); 17 were controls and 60 were technical replicates run either in the same lab (intra-site replicate) or in different labs (inter-site replicate). All cases were double-blinded in this study and divided into several categories: 1) cases with known pathogenic or likely pathogenic variants (n=73 amniotic fluid, n=5 CVS), 2) cases with known variants of uncertain significance (n=27 amniotic fluid), 3) cases with no known reportable variant (n=14 amniotic fluid, n=4 CVS), and 4) genome controls (n=17 presumably healthy donor blood). Only variants reported from SOC methods were considered for concordance comparison.

### Whole genome SV calls and SV filtering

#### Data QC and EnFocus^™^ Fragile X analysis

All 200 datapoints were analyzed using the *de novo* assembly pipeline with approximately 800 Gbp of data used for each dataset. Additionally, all samples were analyzed for the Fragile X repeat expansion using the EnFocus™ Fragile X analysis pipeline as a screen for incidental findings. Quality control metrics were exceptional with the average molecule N50=277 kbp, map rate of 85.2%, and average effective coverage of 209x. The EnFocus™ Fragile X pipeline and whole genome pipeline were used for postanalytical QC provided a pass/fail analysis for the following assessments: 1) inferred sex was determined and reported to be 112 males, and 88 females which was 100% concordant with the accession information (self-reported), 2) stable regions analysis (internal control for assembled map measurement precision) resulted in 100% samples passing (200/200), and 3) assessment of the CNV noise level resulted in 192/200 cases as passed. Despite eight samples failing CNV noise assessment, all samples were analyzed, and these eight were still concordant to SOC testing results. Assessment of the *FMR1* region for the determination of any cases harboring potential full mutation did not result in any positive findings (Table S3).

#### Genome wide SV analysis

The following filters were used: deletions and insertions >1500 bp, CNVs from coverage depth pipeline >500 kbp, present in ≤1% of the control database, and SVs within 3 kbp and CNVs within 500 kbp of a known gene. The average number of SVs per sample was 4,241.7 (min=3,540, max=4,773) from the baseline set of software default filtration criteria described earlier. This further filtering scheme resulted in an average of 16.4 variants per sample to be analyzed and classified by the variant analyst and director.

#### AOH/ROH Detection

Four cases were identified with areas of homozygosity (AOH) by CMA – two of which were instances of consanguinity. Of those four, two cases (BNGOSS-0001344 and BNGOSS-0001369) were >25Mb and detected by OGM. Other AOH were below the size threshold of detection by OGM.

### Concordance of OGM to SOC testing results

A technical concordance of 100% (200/200) was achieved in this study. All SVs and CNVs observed and reported in SOC testing also were detected in this study by OGM, as were AOH >25Mb.

### Replicate analysis (intra-run, inter-run, inter-site)

A total of 32 unique cases were used to generate 83 datapoints to assess the reproducibility of the assay and workflow. The reproducibility of SVs, CNVs and aneusomies on 37 calls was evaluated, and that aggregated to 96 total replicate variants (39 duplications, 17 deletions, three translocations, 30 trisomies and seven monosomies). In total, there was 100% reproducibility among replicates for all SVs, CNVs and aneusomies.

Furthermore, the precision of SV and CNV breakpoints and sizes of the duplication, deletion and translocation calls were assessed and revealed that SVs detected by the SV pipeline have a higher resolution and size agreement compared to those detected by the CNV pipeline (median breakpoint recall precision SD = 30,293 bp, median size variability SD = 143,341 bp) (Table S2 and Figure S1).

### Representative example: OGM consolidates sequential SOC testing

BNGOSS-0000636 case highlights the utility of OGM. SOC CMA testing detected a 43.1 Mbp gain of chromosome 2q31.2q36.1. However, given the size and interstitial nature of this copy number gain, KT studies were performed to evaluate whether this copy number variant represented a structural rearrangement. While KT identified additional material on chromosome 14, the chromosome morphology was insufficient to definitively conclude whether this represented the variant detected by CMA studies. As such, dual color BAC FISH analysis was performed to confirm the localization and determination of the orientation of the 2q region on chromosome 14. Taken together these studies suggest the prenatal findings were due to an apparent unbalanced der(14)t(2;14). Despite three different methods employed during routine SOC testing, the insertion point on chromosome 14 and whether any genes were disrupted could not be determined. Figure 3 shows that OGM readily detected the presence, location and orientation of the 2q gain, and revealed that the 2q region was, in fact, inserted in an inverted orientation into an interstitial region of chromosome 14 in a manner that disrupted the *JAG2* gene. In this study, 56% of cases (69/123) had two SOC tests performed to reach a diagnosis and 19% of cases (23/123) required three different SOC techniques (Supplementary table 1).

**Figure 3.**
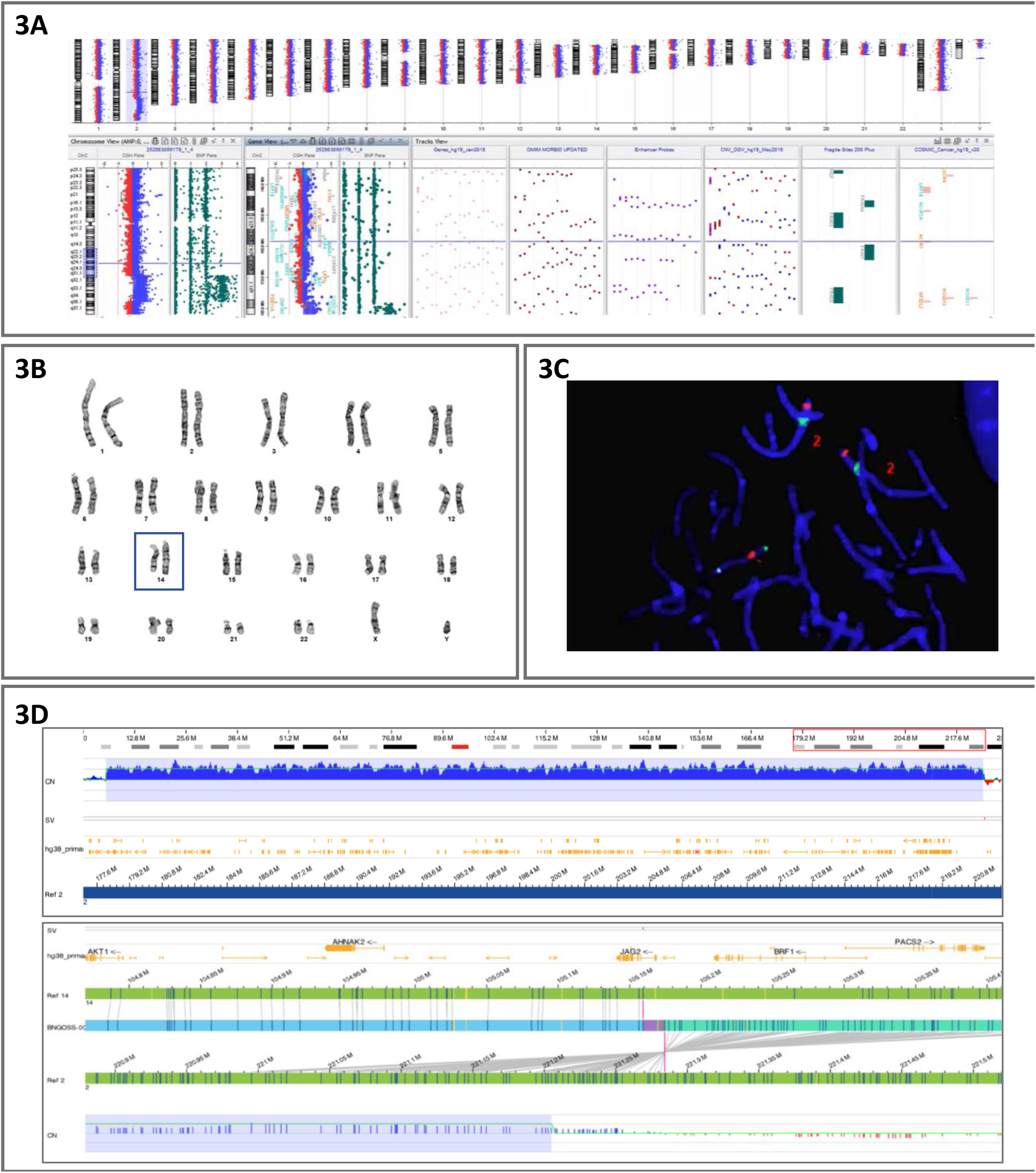
Case BNGOSS-0000636 where multiple SOC methods were used for SV detection. 3A) CMA results from sample show a large duplication on Chr2q. 3B) Karyotyping shows an apparently unbalanced translocation involving chromosomes 2 and 14. 3C) Dual color BAC FISH confirmed the karyotying results with ambiguity regarding the location of the duplication. 3D) OGM results showing a 43.1 Mbp gain of 2q31.2q36.1 inserted interstitially in an inverted orientation into 14q32.33 disrupting the JAG2 gene.

## Discussion

The first phase of this multi-site prenatal study concentrated on concordance and reproducibility of OGM analysis compared to SOC methodologies. In line with multiple published studies, the overall technical concordance was 100% (17–19). One clear advantage of OGM analysis highlighted in this study is its ability to detect and characterize all classes of CNVs and SVs, and screen for genome-wide repeat expansions >500 bp, in a single platform. A unique benefit of OGM is the ability to run an additional analysis pipeline for the screening of individuals with an expanded allele in the *FMR1* gene that could be causative of FXS. Currently, the FXS testing is performed as a separate SOC test (24).

An illustrative example of how OGM resolved multiple sequential tests in one assay is shown by case BNGOSS-0000636 (Figure 3). The ability to quickly determine the complexity of genomic aberrations is critical for prenatal diagnostic testing. As demonstrated by this case, three different methods were initially required (due to each having its own limitations) to shed light on the SV being an unbalanced derivative chromosome 14 [der(14)] resulting from an ins(14;2). Clearly, the CMA and KT results allowed for determining the variants to be pathogenic and recommended further parental follow-up testing. It is important to note that *JAG2* is associated with autosomal recessive limb-girdle muscular dystrophy (OMIM 619566). In the event of a balanced parental carrier of the insertion, carrier testing of the other parent for a *JAG2* variant may be warranted. This case exemplifies the value of using a technique that is more comprehensive and sensitive such that tiered testing is no longer essential or needed less often. OGM shortens the time to actionable results which is critical in the prenatal testing to make informed reproductive and clinical management decisions. Indeed, 75% of the cases in this study were tested using at least two different cytogenetic assays, and 19% of cases had CMA, FISH and KT performed (Figure 1A). Specifically, this example shows that OGM has the potential to become a first line test as it would have likely reduced the time to attain comprehensive answers at a reduced cost.

In addition to the reduced TAT and cost saving benefits to patients, OGM also offers workflow benefits to laboratories. Routine cytogenetic analysis requires experienced technologists that spend a significant amount of time performing microscopic analysis and karyotyping metaphases. Additional techniques such as FISH and CMA require further laboratory personnel and equipment, adding extra time to perform these assays. Cytogenetic laboratory staffing issues, fueled by diminished technologist training programs, are steadily impacting the clinical practice of cytogenetic testing on a global scale (unpublished data, (25,26). Since these personnel shortage issues are only expected to worsen, it is imperative to evaluate modalities of testing that take into account the present challenges affecting clinical cytogenetic laboratories. The end-to-end complete solution offered by OGM addresses the analysis and interpretive aspect of cytogenomic testing where the Bionano Access software addresses the needs for the SVs to be analyzed and interpreted by the laboratory staff with minimal training and experience.

## Conclusions

OGM has the potential to significantly impact cytogenomic diagnostics in the prenatal setting by offering the combined diagnostic power of KT, FISH and CMA in a single assay. In addition to detecting large scale copy number changes and submicroscopic CNVs, OGM also may uncover balanced structural rearrangements and characterize their specific architecture, highlighting disrupted genes at the breakpoint regions. Although not described in this study, a similar study on postnatal disorders has demonstrated that OGM has the ability to detect specific repeat expansion signatures such as seen in the Fragile X syndrome (accepted for publication in the Journal of Molecular Diagnostics, Dec., 2022). The standardized workflow and cost-effectiveness of OGM coupled with its ability to provide high resolution cytogenomic analysis in a single assay, indicate that OGM has the potential to serve as a first-tier diagnostic test for prenatal diagnosis.

IRB and consent disclosures:

IRB-00007527 – University of Rochester Medical Center Office for Human Subject Protection

IRB-AAAT9083 – Columbia University Irving Medical Center, New York, NY, USA

IRB-2022-0865 – Cincinnati Children’s Hospital Institutional Review Board, Cincinnati, OH, USA

IRB-20212956 – Bionano Genomics Inc., San Diego, CA, USA

IRB-22-37290 – University of California San Francisco, San Francisco, CA, USA

IRB-A-#00000150 (HAC IRB # 611298) - Medical College of Georgia, Augusta University, Augusta, GA, USA

## Data Availability

anonymized data may be made available upon request

## Acknowledgements

The authors wish to acknowledge Bionano clinical affairs team: Alka Chaubey, Alex Hastie, Rachel Burnside, Mike Gallagher, James Yu, Ben Clifford, Vruti Mehta, Andy Pang, Alex Chitsazan, Joey Estabrook, Kelsea Chang for support in training and data management.

