## Supplementary figures for "Multisite evaluation and validation of Optical Genome Mapping for prenatal genetic testing"

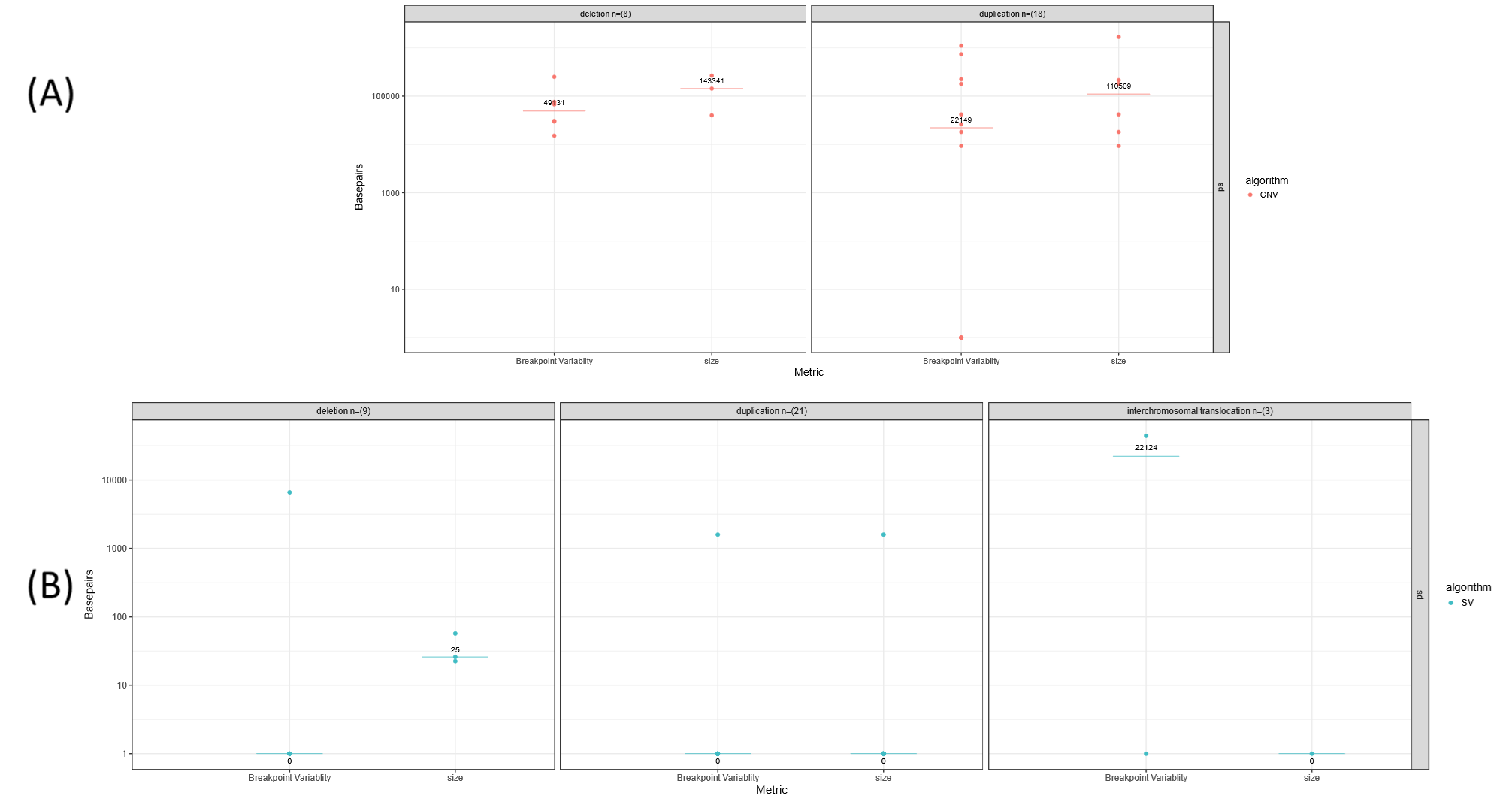

**Supplementary Figure 1: Breakpoint variability and size agreement:** (A.) Median breakpoint variability of position 1 and position 2 for deletions (Breakpoints = 16, number of variants = 8, cases = 2) , and duplications (Breakpoints = 36, number of variants = 18, cases =6) of the SV algorithm (B) Breakpoint variability of position 1 and position 2 for deletions (Breakpoints = 18, number of variants = 9, cases = 3) and duplications (Breakpoints = 42, number of variants = 21, cases = 9), and intrachromosomal translocations (Breakpoints = 6, number of variants = 3, cases = 1) of the CNV algorithm

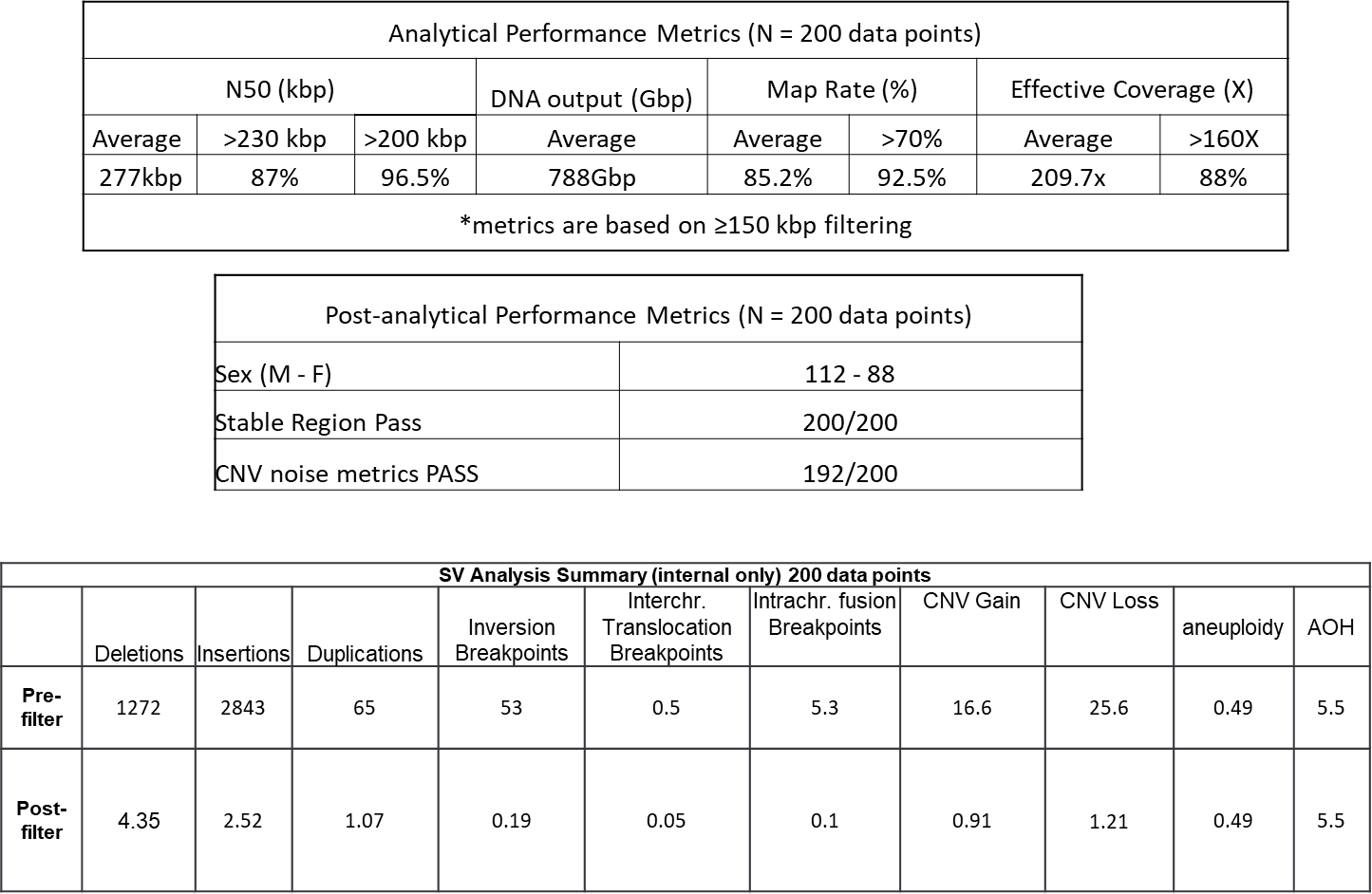

**Supplementary Figure 2:** QC and variant calling. The results of analytical and post-analytical QC metrics as defined in the Methods section are shown in the first and second tables. The third table shows the average number of SV counts by type before and after the SV filtering steps. An inversion breakpoint involves neighboring alignments with opposite orientations. A translocation (or fusion) point between distant regions of the genome is identified as a translocation breakpoint. Intrachromosomal fusion breakpoints involve regions typically at least 5 Mbp away from each other on the same chromosome, whereas interchromosomal translocation breakpoints involve regions on different chromosomes. (reference: https://bionanogenomics.com/wp-content/uploads/2018/04/30110-Bionano-Solve-Theory-of-Operation-Structural-Variant-Calling.pdf)
